# Educational Preparation, Social Media Use, and Ethical Decision-Making in Healthcare Delivery: A Qualitative Case Study from Ghana

**DOI:** 10.64898/2026.01.08.26343746

**Authors:** Charles Owusu-Aduomi Botchwey, Lawrencia Aggrey-Bluwey, Bernice Ohenewaa Asare, Johnson Sam, Peter Bakela Ndewini, Emmanuel Sei Nkpetri

## Abstract

**Objective:** This study examined how educational preparation and social media use shape healthcare delivery among healthcare professionals in a district-level public hospital in Ghana. It explored how education influences health communication and patient engagement, how social media platforms are used in routine practice, and how ethical concerns shape digital engagement.

**Study Design:** An exploratory qualitative case study was conducted within an interpretivist paradigm to capture healthcare professionals’ experiences and interpretations of digital communication in everyday clinical practice.

**Methods:** Thirty healthcare professionals from different cadres were purposively sampled from a district-level public hospital. Data were collected through semi-structured interviews conducted between May and July 2024. Interviews were audio-recorded, transcribed verbatim, and analyzed thematically using Braun and Clarke’s six-step approach. A hybrid deductive and inductive coding strategy was applied, informed by sensitizing concepts from the Technology Acceptance Model and the Health Belief Model.

**Results:** Participants described strong confidence in their medical knowledge and communication skills, shaped by formal education and continuous learning, which supported both face-to-face and digital patient engagement. Social media use was selective and purpose-driven. WhatsApp was commonly used for patient reminders and follow-up communication, Facebook for community health outreach, and Twitter for professional learning. Decisions to engage digitally were mediated by concerns about misinformation, confidentiality, and professional boundaries. In the absence of clear institutional guidance, healthcare professionals relied on personal judgment to balance the perceived usefulness of social media with ethical risk, resulting in cautious and bounded digital practices.

**Conclusion:** The findings show that healthcare professionals’ use of social media is shaped by the interaction of education, perceived usefulness, ethical responsibility, and institutional context. By integrating the Technology Acceptance Model and the Health Belief Model, the study demonstrates that digital engagement in healthcare involves both adoption and restraint. The study provides context-specific insight from a district-level hospital and highlights the need for clear policies, training, and institutional support to promote safe and effective digital communication in healthcare.

## Introduction

The use of digital platforms in healthcare delivery has expanded rapidly over the past decade, particularly in low- and middle-income countries where mobile phone access has outpaced the development of formal digital health systems [1,2]. Globally, more than 5 billion people now use mobile phones, and over 60 percent of adults report seeking health information online at least occasionally [3,4]. In sub-Saharan Africa, mobile broadband subscriptions increased by more than 40 percent between 2018 and 2023, creating new pathways for health communication outside formal clinical settings [5].

In Ghana, digital connectivity has grown steadily. Recent national data indicate that over 70 percent of adults own a smartphone, and WhatsApp and Facebook remain among the most widely used platforms across age groups [6,7]. The Ghana Health Service has acknowledged that social media has become an important channel for public health messaging, patient follow-up, and community engagement, particularly during public health emergencies such as COVID-19 [8]. At the same time, national reports have highlighted persistent challenges related to misinformation, weak regulation of online health content, and uncertainty about professional boundaries in digital communication [9,10].

Healthcare professionals play a critical role in this digital environment. Studies across Ghana and similar contexts show that nurses, midwives, physician assistants, and public health practitioners are increasingly approached by patients for clarification of health information encountered online [11,12]. Messaging platforms are often used informally for appointment reminders, medication follow-up, and health education, especially in district-level facilities where patient volumes are high and continuity of care can be difficult to maintain [13]. Despite this growing reliance on digital tools, formal institutional guidance on professional social media use remains limited in many public healthcare facilities [9,14].

Existing literature on social media and healthcare has documented both opportunities and risks. On one hand, digital platforms have been shown to support patient engagement, expand access to health information, and enhance health promotion activities [2,4,15]. On the other hand, research consistently points to the spread of misinformation, breaches of confidentiality, and blurred professional roles as significant concerns [10,16,17]. In Ghana, empirical studies suggest that healthcare professionals are aware of these risks and often adopt cautious digital practices, yet these practices are shaped largely by individual judgment rather than formal policy [11,12].

Importantly, much of the existing research treats education, technology use, and ethics as separate domains. Studies on digital health adoption often focus on access and skills, while ethical discussions are addressed in isolation, and professional education is examined primarily in relation to clinical competence [15,18]. This fragmented approach limits understanding of how these elements intersect in everyday practice, particularly in district-level hospitals where frontline workers must balance patient needs, institutional constraints, and ethical responsibility with minimal organisational support.

District-level hospitals represent a critical but under-researched setting in Ghana’s health system. These facilities deliver essential services to large populations and often serve as the first point of referral for surrounding communities [19]. Healthcare professionals working in these settings face high workloads and resource constraints, which may increase reliance on informal digital communication. Yet evidence on how these professionals navigate social media use in relation to their training and ethical obligations remains sparse [11,14].

This study addresses this gap by examining how educational preparation and social media use shape healthcare delivery among healthcare professionals working in a district-level public hospital in Ghana. It explores how education influences health communication and patient engagement, how social media platforms are used to disseminate and verify health information, and how ethical concerns shape digital practice. These insights are particularly relevant for informing digital health policy and professional guidance in settings where technological adoption continues to outpace regulation [9,10].

## Theoretical Underpinning

This study is grounded in two complementary theoretical frameworks. These are the Technology Acceptance Model (TAM) and the Health Belief Model (HBM). Together, these frameworks provide an analytical lens for understanding how healthcare professionals adopt digital tools while simultaneously exercising caution in their use. The combined application of these models is particularly relevant in contexts where digital technologies are widely available but institutional guidance remains limited [11,15].

TAM has been widely used to explain individuals’ adoption of information technologies across healthcare settings [16,17]. In health research, TAM has been applied to examine acceptance of electronic health records, telemedicine platforms, and mobile health applications [17,20]. The model focuses on how perceived usefulness and perceived ease of use shape individuals’ willingness to engage with technology. In the context of this study, TAM provides a useful framework for examining why healthcare professionals perceive certain social media platforms as appropriate for patient communication, follow-up, and health education. It helps explain platform preferences, frequency of use, and the integration of digital tools into routine practice.

However, TAM alone offers limited insight into why healthcare professionals may deliberately restrict or avoid certain forms of digital engagement despite recognizing their functional benefits [15,21]. In healthcare contexts, technology use is closely tied to ethical responsibility, professional identity, and perceptions of risk. These dimensions extend beyond technical acceptance and require a behavioral framework that accounts for judgment and restraint. HBM complements TAM by focusing on how perceptions of risk and responsibility shape behavior [14,18]. HBM has been extensively applied in health research to explain decision-making related to prevention, treatment adherence, and professional practice [18,22]. Key constructs of the model include perceived severity, perceived susceptibility, perceived barriers, self-efficacy, and cues to action. In the context of digital communication, these constructs are particularly relevant for understanding how healthcare professionals assess the potential consequences of sharing information online, manage concerns about confidentiality, and decide when digital engagement is appropriate.

In this study, HBM provides a framework for interpreting healthcare professionals’ cautious and selective use of social media. Perceived severity and perceived barriers help explain concerns about misinformation, misinterpretation, and breaches of confidentiality. Self-efficacy reflects confidence in professional knowledge and communication skills, which supports engagement when risks are considered manageable. The absence of institutional guidelines can be understood as a lack of formal cues to action, which contributes to uncertainty and variability in practice [9,14].

The integration of TAM and HBM allows for a more comprehensive understanding of digital engagement among healthcare professionals than either framework alone. TAM explains why social media platforms are perceived as useful and accessible tools for patient communication.

HBM explains why these same platforms are used cautiously and within clearly defined boundaries. Together, the frameworks capture both adoption and restraint as interconnected processes rather than opposing outcomes [15,21].

## Methodology

### Study Design

This study followed the Standards for Reporting Qualitative Research as outlined by O’Brien et al. [19]. An exploratory qualitative case study design was adopted within an interpretivist paradigm, which recognizes that meaning is constructed through individuals’ lived experiences and social contexts [18]. The case study approach was selected to enable an in-depth examination of how healthcare professionals working within a single institutional setting interpret their educational preparation, engage with social media, and navigate ethical concerns in everyday practice. Focusing on one hospital allowed for close attention to contextual influences, organisational routines, and shared professional norms that shape digital engagement.

### Study Setting

The study was conducted at Ahmadiyya Islamic Hospital, a district-level public hospital located in the Agona West Municipality of Ghana. The facility provides outpatient, inpatient, maternal, and community-based services to a mixed urban and peri-urban population. The hospital plays a central role in the local health system and serves as a first point of referral for surrounding communities. Preliminary engagement with staff indicated that social media platforms were commonly used for routine communication, patient follow-up, and dissemination of health information. This made the hospital an appropriate and information-rich case for examining digital communication within everyday healthcare delivery.

### Participants and Sampling

Participants were healthcare professionals who had worked at the hospital for at least one year and who reported active use of at least one social media platform for personal or professional purposes. The sample included registered general nurses, midwives, physician assistants, public health nurses, and health information officers. Given this diversity of professional roles, a maximum-variation purposive sampling strategy was employed [20]. This approach was intentional and aimed to capture a wide range of perspectives on how education, digital engagement, and ethical considerations intersect across different cadres within the same institutional context.

A total of thirty participants were recruited and interviewed. Data saturation was reached after approximately twenty-five interviews, at which point no substantively new insights were emerging. The remaining interviews were conducted to confirm the stability and completeness of the developing themes and to ensure adequate representation across professional roles.

### Data Collection Instrument

Data were collected using a semi-structured interview guide developed in alignment with the study objectives, relevant empirical literature [7,15,16], and concepts drawn from TAM and HBM. Constructs from TAM informed questions related to perceived usefulness and perceived ease of use of social media platforms [16]. Constructs from HBM informed questions related to perceived benefits, perceived barriers, perceived severity, and professional self-efficacy [14].

The interview guide explored participants’ educational preparation, communication practices, use of social media in patient engagement, approaches to verifying health information, and experiences of ethical uncertainty. Open-ended questions encouraged participants to reflect on concrete examples from their daily work rather than abstract opinions. The guide was piloted with two healthcare professionals from a different district-level facility to assess clarity and flow. Minor revisions were made following the pilot to strengthen probing around ethical dilemmas and platform-specific decision-making.

### Data Collection Procedures

Data were collected between May 1 and July 2, 2024. Face-to-face interviews were conducted in English in quiet locations within the hospital to ensure privacy and minimize disruption to clinical duties. Interviews lasted between thirty and sixty minutes. All interviews were audio-recorded with participants’ consent to ensure accurate capture of their accounts. Field notes were taken during and immediately after each interview to document contextual details, non-verbal cues, and initial analytical reflections. These notes were later used to enrich interpretation during analysis.

### Data Analysis

Data analysis followed Braun and Clarke’s six-step approach to thematic analysis [21]. Analysis began with repeated reading of the transcripts to achieve familiarization with the data. Coding was conducted using a hybrid deductive and inductive strategy. Deductive codes were informed by the study objectives and key constructs from TAM and HBM, which were used as sensitizing concepts rather than rigid categories. At the same time, inductive codes were generated directly from participants’ accounts to allow unanticipated patterns and meanings to emerge. Codes were iteratively reviewed and refined through constant comparison across transcripts. Related codes were clustered into preliminary themes, which were then examined for coherence, distinctiveness, and alignment with both the data and the theoretical underpinnings of the study. Regular analytic discussions were held among members of the research team to compare interpretations, refine theme boundaries, and enhance analytic depth. Microsoft Word and Excel were used to organize transcripts, coding matrices, and analytic memos. An audit trail documenting analytic decisions was maintained throughout the process.

### Trustworthiness

Trustworthiness was addressed in line with guidance by Creswell and Poth [18]. Credibility was enhanced through member checking, whereby participants reviewed summaries of their interviews to confirm accuracy and meaning. Field notes were used to triangulate interview data and support interpretation. Transferability was supported through detailed description of the study setting, participants, and analytic process, enabling readers to assess relevance to other contexts. Dependability was ensured through transparent documentation of sampling decisions, interview procedures, and analytic steps. Confirmability was strengthened through reflexive journaling, which allowed the researchers to examine how their professional backgrounds and assumptions could influence interpretation.

### Ethical Considerations

Ethical approval for the study was obtained from the University of Education, Winneba Ethical Review Board, with clearance number UEW-ERB/HS/2024/018. Written permission was also granted by the hospital administration. All participants provided informed consent after receiving detailed information about the study purpose and procedures. Participation was voluntary, and participants could withdraw at any time without consequence. Measures were taken to protect anonymity and confidentiality. Audio recordings and transcripts were stored securely, and no identifying information was included in the report.

## Findings

The findings describe how healthcare professionals at Ahmadiyya Islamic Hospital interpret their educational preparation, use social media in everyday practice, and manage ethical uncertainty in digital communication. The analysis followed a hybrid deductive and inductive approach. Sensitizing concepts from TAM and HBM informed early coding. Themes were refined through repeated engagement with the data. The final structure reflects how theoretical ideas were expressed through routine professional decisions within a district-level hospital context.

### Theme 1: Educational Preparation and Professional Confidence in Communication

Participants consistently linked their communication practices to their educational background and ongoing professional learning. Education was described as more than formal qualification. It was framed as a process that built confidence, shaped judgment, and defined professional boundaries.

Participants repeatedly emphasized their responsibility to translate medical knowledge into language patients could understand. Several participants described confidence as emerging through training and reinforced by practice.

One registered general nurse explained, “*As a nurse, I’ve been trained to explain medical issues in ways that patients can understand. Whether it is in the ward or on WhatsApp, I am confident in what I say because I know I have been trained to do this well*” - Participant 4.

This account illustrates how formal education supported communication across both physical and digital settings. Other participants emphasized the role of continuous learning. A public health nurse described how staying updated reduced uncertainty during online engagement.

“*I read and update myself regularly, so I am not worried when people ask me about medical issues on Facebook or in our health group. I feel prepared and sure of the information I share*” - Participant 8.

For this participant, ongoing education strengthened confidence and supported willingness to engage digitally. Participants also described limits to this confidence. A midwife explained how she exercised caution when responding to online health queries.

“*Sometimes patients send me messages online asking about their symptoms. I take my time to respond. I only give answers I am sure about. If I am not certain, I refer them*” - Participant 11.

These accounts address the first study objective by showing how education contributes to health literacy and patient engagement. Education strengthened professional self-efficacy, a core construct of the HBM. Participants believed they were capable of communicating accurate information. At the same time, awareness of potential harm moderated their actions. Digital engagement was therefore shaped by confidence and by perceived severity. From a TAM perspective, confidence increased perceived usefulness of social media for patient communication, while caution limited the scope of use.

### Theme 2: Negotiating Comfort in Provider-Patient Communication Across Settings

Participants described differences in comfort between face-to-face communication and digital interaction. In-person encounters were described as familiar and structured. Digital communication introduced new dynamics that required adjustment. Comfort depended on perceived clarity, feedback, and professional control. Some participants viewed digital communication as supportive of patient engagement.

One nurse explained, “*I feel relaxed when chatting with patients through WhatsApp. It is easier to explain things when people are not rushed or nervous like they are in the hospital*” - Participant 2.

This environment was perceived as less intimidating for patients and more flexible for providers. Other participants expressed discomfort with online exchanges.

A physician assistant explained, “*I prefer face-to-face interactions. Online communication can be misunderstood. I want to see the person’s reaction. It helps me know if they understand*” - Participant 14.

For this participant, the absence of visual cues increased uncertainty and reduced confidence in digital communication. Several participants also described adopting selective engagement strategies.

A midwife explained, “*I reply to patients’ messages when it is something simple. I avoid giving too much advice online. Anything complicated, I ask them to come in*” - Participant 9.

These findings further address the first objective by illustrating how communication comfort shapes patient engagement. Differences in comfort reflect perceived barriers and perceived severity within the HBM. Digital communication was adopted when it felt manageable and restrained when risks were perceived as high. From the TAM perspective, perceived ease of use influenced willingness to engage. When platforms supported clarity and control, they were used more readily.

### Theme 3: Trust, Verification, and the Management of Health Information Online

Participants expressed strong concern about the quality of health information circulating on social media. Misinformation was described as widespread and dangerous. Professional training shaped how participants evaluated credibility and decided what to share. A public health nurse described her criteria for trust:

“*I do not trust health tips on social media unless they come from WHO, Ghana Health Service, or a medical journal. There is too much false information online”* - Participant 1. Credibility was therefore linked to institutional authority rather than online popularity.

Other participants described verification as a routine professional practice. One nurse explained, “*WhatsApp groups share all kinds of health tips. Before I forward anything, I double-check. I search online or ask colleagues*” - Participant 6.

Verification was framed as part of professional responsibility. First-hand experiences reinforced caution. One participant explained, “*I have seen people use herbal remedies they found on Facebook and end up with complications*” - Participant 12.

These experiences increased vigilance and skepticism. These findings address the second study objective by explaining how healthcare professionals engage with digital information dissemination. Participants’ actions reflect perceived susceptibility and perceived severity within the HBM. Misinformation was seen as likely and harmful. These perceptions shaped cautious behavior. From the TAM perspective, credibility influenced perceived usefulness. Social media was used as an information channel only when trust could be established.

### Theme 4: Platform Use as Strategic and Context-Dependent Practice

Participants described platform choice as intentional and situational. Different platforms served different purposes. Decisions were shaped by accessibility, audience, and risk. WhatsApp was described as the most practical platform.

One nurse explained, “*I use WhatsApp every day. We have patient groups and community groups. It is the fastest way to communicate*” - Participant 3.

Familiarity and widespread use supported routine engagement. Facebook was used for outreach rather than personal communication.

A health information officer explained, “*Facebook is where I share health posts during outreach programs. It reaches many people*” - Participant 7.

Privacy concerns however limited its use for individual patient interaction. Twitter was described as a professional space.

A physician assistant explained, “*Twitter gives me updates from global health organizations. I rarely interact with patients there*” - Participant 10.

This platform supported professional learning rather than patient care. These findings address the second objective by showing how platform preferences shape information dissemination. Choices reflect perceived usefulness and perceived ease of use from the TAM. Social norms and patient access also influenced decisions. The absence of institutional guidance meant that platform use depended on individual judgment and experience.

### Theme 5: Ethical Uncertainty and the Absence of Institutional Guidance

Ethical concerns featured prominently across participants’ accounts. Confidentiality was described as a persistent source of anxiety. Digital communication was perceived as difficult to control once information was shared.

A midwife explained, “*I worry about privacy. If something is shared out of context, it can cause problems*” - Participant 5.

Participants repeatedly highlighted the lack of formal guidance. *A nurse explained, “We do not have clear guidelines from management. It is trial and error*” - Participant 13. Without institutional policies, professionals relied on personal judgment.

Despite these concerns, participants continued to use social media for patient education. One nurse described sending reminders. “*I send messages about medication or blood pressure management. It helps patients remember*” - Participant 15. Others emphasized using verified materials. “*I use flyers or infographics from health authorities. Otherwise, people will not trust it*” - Participant 6.

These findings address the third study objective by highlighting ethical considerations associated with digital engagement. Participants’ concerns reflect perceived severity and perceived barriers within the HBM. The absence of institutional cues limited confidence in digital practice. From the TAM perspective, lack of organisational support constrained adoption even when perceived usefulness was high.

## Discussion

This study examined how educational preparation, social media use, and ethical considerations shape healthcare delivery among professionals working in a district-level hospital in Ghana. The findings show that digital engagement is neither automatic nor uniform. Instead, it is shaped by professional training, perceptions of risk, and the institutional environment within which healthcare workers operate. By combining the TAM and the Health Belief Model, the study provides a layered explanation of how healthcare professionals adopt digital tools while simultaneously exercising restraint.

### Education as the Foundation of Digital Confidence and Professional Judgment

A central contribution of this study is the demonstration of how education functions as the foundation of digital engagement rather than simply as background context. Participants repeatedly linked their ability to communicate with patients to formal training and continuous learning. Education shaped not only what participants knew, but how confident they felt in sharing information and how they judged the limits of appropriate communication. This finding aligns with earlier work showing that provider confidence is critical for effective patient engagement and health communication [5]. However, the present study extends this literature by showing how educational confidence operates within digital environments.

From the perspective of the HBM, participants’ accounts strongly reflect the construct of self-efficacy [14]. Healthcare professionals believed they were capable of explaining medical information clearly and responsibly. This belief supported engagement through platforms such as WhatsApp and Facebook. At the same time, education informed professional restraint. Participants did not describe digital engagement as unrestricted. Instead, they demonstrated awareness of potential harm, which reflects perceived severity within the HBM. This balance between confidence and caution suggests that self-efficacy does not automatically lead to increased digital activity. Rather, it interacts with risk assessment to shape practice.

These findings complement studies conducted among nurses in rural Ghana, which found that professional training influenced willingness to adopt social media for health education [6]. Unlike some prior studies that emphasize technology skills alone, the present findings highlight education as a moral and professional anchor that guides digital behavior. This distinction is important in low-resource settings where formal digital health guidelines are still emerging.

### Communication Comfort and the Limits of Digital Interaction

The findings also show that comfort with patient communication varies across settings. Face-to-face encounters were described as structured and predictable. Digital communication required adaptation and introduced uncertainty for some participants. This pattern reflects earlier research showing that healthcare professionals value visual cues and immediate feedback in clinical communication [16]. In the present study, the absence of these cues shaped cautious engagement for some participants.

From the HBM perspective, discomfort with digital communication reflects perceived barriers [14]. Fear of misunderstanding or misinterpretation reduced willingness to engage fully online. From the TAM perspective, perceived ease of use influenced adoption [11]. Platforms that felt intuitive and manageable were more readily used. Those perceived as ambiguous or risky were restricted to limited functions. Importantly, participants did not frame digital communication as replacing face-to-face care. Instead, they described it as complementary. This finding contrasts with some digital health narratives that suggest technology will transform care delivery entirely. Instead, the study supports more recent qualitative work showing that healthcare professionals integrate digital tools selectively while preserving traditional clinical boundaries [4]. This selective integration reflects professional norms rather than technological resistance.

### Misinformation, Verification, and Professional Gatekeeping

Concerns about misinformation were pervasive in participants’ accounts. Healthcare professionals described social media as a space where inaccurate health information circulates rapidly. These concerns are consistent with broader literature on the infodemic and information quality in digital health communication [7]. In the present study, professional training shaped how participants responded to this challenge. Participants described verification as a routine part of their work. This behavior reflects high perceived susceptibility and perceived severity within the Health Belief Model [14]. Participants believed that patients were likely to encounter misinformation and that the consequences could be serious. These beliefs motivated careful filtering of information. From the Technology Acceptance Model perspective, credibility strongly influenced perceived usefulness [11]. Social media platforms were only considered useful when they could support the dissemination of accurate information.

This finding aligns with research showing that Ghanaian health professionals apply selective trust when engaging with digital content [7]. The present study adds depth by showing how verification practices are embedded in everyday routines rather than treated as exceptional tasks. Healthcare professionals positioned themselves as intermediaries who translate digital information into clinically appropriate messages. This gatekeeping role is particularly important in contexts where patients rely heavily on informal digital sources for health advice.

### Platform Choice as Situated and Strategic Practice

The study found that platform use was deliberate and context-dependent. WhatsApp, Facebook, and Twitter served different functions within participants’ professional lives. These choices were shaped by accessibility, audience characteristics, and perceived risk. Similar patterns have been reported in studies of healthcare workers in Cape Coast and other parts of Ghana [4]. However, the present study shows how these patterns operate within a single institutional setting. From the TAM perspective, platform choice reflects perceived usefulness and perceived ease of use [11]. WhatsApp was favored because it aligned with patients’ communication habits and required minimal technical effort. Facebook supported broader outreach. Twitter functioned as a professional learning space rather than a patient-facing tool. These distinctions demonstrate that social media use among healthcare professionals is not uniform. It is shaped by context and purpose.

The absence of institutional guidance amplified the role of individual judgment. Without formal policies, healthcare professionals developed their own strategies for platform use. This finding resonates with earlier research highlighting institutional unpreparedness for regulating digital health technologies in Ghana [9]. It also explains variation in practice within the same hospital. Digital engagement became a negotiated practice rather than a standardized organisational process.

### Ethical Uncertainty and the Consequences of Institutional Silence

Ethical concerns emerged as a central theme across participants’ narratives. Confidentiality was a persistent worry. Participants expressed uncertainty about boundaries and responsibility in digital spaces. These concerns are widely documented in studies examining social media use among healthcare professionals [16]. In the present study, ethical uncertainty was intensified by the absence of formal institutional guidance. Within the HBM, ethical concerns reflect perceived severity and perceived barriers [14]. Fear of breaching confidentiality discouraged unrestricted digital engagement. The absence of institutional cues limited confidence in decision-making. From the TAM, lack of organisational support constrained adoption even when perceived usefulness was high [11].

Despite these constraints, participants continued to use social media for patient education. They relied on verified materials and limited the scope of digital interaction. This behavior suggests active negotiation rather than disengagement. Healthcare professionals balanced professional obligation with ethical caution. This finding extends prior work by showing how ethical uncertainty shapes daily practice rather than abstract attitudes [9].

### Integrating TAM and HBM to Explain Digital Engagement

The combined use of TAM and HBM offers a more comprehensive explanation of healthcare professionals’ digital behavior than either model alone. TAM explains why certain platforms are adopted based on usefulness and ease of use [11]. HBM explains why adoption is cautious, selective, and ethically constrained [14]. Education strengthened self-efficacy and increased perceived usefulness. Misinformation and confidentiality concerns increased perceived severity and barriers. The absence of institutional guidance limited cues to action. These factors interacted to shape digital engagement as a negotiated process. This integrated approach responds to calls in the literature for multi-theoretical frameworks that capture both technological and behavioral dimensions of digital health practice [15].

#### Contribution to Digital Health Research

This study contributes to digital health research by providing context-specific insight from a district-level hospital in Ghana. Much existing research focuses on urban or policy-level settings [4,7]. By examining frontline practice, the study shows how healthcare professionals integrate digital tools under conditions of limited guidance and high responsibility. The findings demonstrate that healthcare professionals are reflective practitioners who adapt technology to align with training, ethics, and patient needs. This contribution is particularly relevant for low-resource settings where digital adoption often outpaces regulation. The study therefore adds empirical depth to ongoing debates about responsible digital health engagement in sub-Saharan Africa.

### Conclusion, Policy Implications and Recommendations

This study examined how healthcare professionals in a district-level hospital in Ghana understand and enact digital communication in their everyday work. The study focuses on educational preparation, social media use, and ethical concerns, and shows that digital engagement in healthcare is shaped by professional judgment rather than by technology alone. Healthcare workers did not describe social media as a neutral tool. They described it as something that required careful assessment, boundary setting, and ethical reflection.

The findings of this study have important implications for health policy and practice in Ghana and in similar settings where digital communication is becoming embedded in everyday healthcare delivery. Healthcare professionals are already using social media to support patient education and follow-up. However, they are doing so in the absence of clear institutional guidance. This gap places the burden of ethical decision-making on individual workers and increases uncertainty in practice.

Health authorities and healthcare institutions should prioritize the development of clear and practical guidelines on professional social media use. Such guidelines should address confidentiality, documentation, patient-provider boundaries, and appropriate scope of digital communication. Policies should recognize that social media is already part of routine care rather than treating it as an optional or future tool. Clear guidance would provide healthcare workers with institutional cues to action and reduce reliance on trial-and-error decision-making.

Training and capacity building are also essential. Digital literacy and ethical communication should be integrated into both pre-service and in-service training programs. These trainings should move beyond technical skills and focus on professional judgment, risk assessment, and responsible information sharing. Strengthening self-efficacy through structured training aligns with the Health Belief Model and supports confident yet cautious digital engagement [14].

Institutions should also invest in the provision of verified digital health materials. Participants in this study relied heavily on trusted sources such as the Ghana Health Service and the World Health Organization when sharing information online. Providing healthcare workers with approved infographics, short educational messages, and visual materials would reduce dependence on unverified sources and improve consistency in patient education. This approach enhances perceived usefulness while reducing ethical risk, which supports sustainable adoption as described by the TAM [11].

At a broader level, policymakers should recognize that digital health adoption in low-resource settings often outpaces regulation. Policies must therefore be responsive to frontline realities. Engaging healthcare professionals in the development of digital communication guidelines can ensure that policies reflect everyday practice rather than abstract ideals. Such engagement would strengthen ownership, improve compliance, and support responsible digital health integration.

### Study Limitations

This study has some limitations that should be considered when interpreting the findings. The research was conducted in a single district-level hospital, which limits the extent to which the findings can be transferred to other healthcare settings. While the case-study approach allowed for in-depth exploration of professional experiences within a specific institutional context, digital communication practices may differ in tertiary hospitals, private facilities, or rural clinics with different resource profiles.

The study also relied on self-reported experiences from healthcare professionals. Participants may have described their practices in ways that reflect professional norms or ethical expectations. This may have influenced how openly challenges and uncertainties were discussed. Although the use of in-depth interviews and reflexive analysis helped to mitigate this risk, the findings remain shaped by participants’ interpretations of their own behavior.

Further, the study focused on healthcare professionals’ perspectives and did not include patients’ views. As a result, the findings reflect how providers understand digital engagement rather than how patients experience or interpret these interactions. Including patient perspectives could provide a more complete picture of how digital communication influences healthcare delivery.

Lastly, the study was conducted in a context where formal digital health policies and institutional guidelines are still evolving. Practices described by participants may change as national or organisational regulations become clearer. This limits the ability to draw conclusions about long-term patterns of digital engagement. However, the findings remain valuable for understanding current practices during a period of transition.

## Declarations

### Ethics Approval and Consent to Participate

Ethical approval for this study was obtained from the University of Education, Winneba Ethical Review Board UEW-ERB/HS/2024/018. Written permission was also granted by the management of the study hospital. All participants received detailed information about the study and provided written informed consent prior to participation. Participation was voluntary, and participants could withdraw at any point without any consequences.

### Consent for Publication

Not applicable. No identifying information of participants is included in this manuscript.

### Availability of Data and Materials

The qualitative interview data generated and analyzed during this study are not publicly available due to ethical and confidentiality considerations. The study was conducted in a single district-level hospital, and the interview transcripts contain detailed contextual information that could allow participants to be identified, even if anonymized. Public sharing of full transcripts was not included in the informed consent process and is restricted by the conditions of ethical approval.

De-identified data extracts supporting the findings of this study are included within the manuscript. Further information may be made available from the corresponding author upon reasonable request, subject to approval by the relevant ethical review board.

## Author Contributions

C.O.-A.B.: Conceptualization, methodology, data collection, formal analysis, writing – original draft.

L.A.-B.: Conceptualization, methodology, supervision, formal analysis, writing – review and editing, project administration.

B.O.A.: Data collection, transcription, data curation, preliminary analysis, writing – review and editing.

J.S.: Methodological input, data interpretation, writing – review and editing. P.B.N.: Data collection, transcription, data curation, writing – review and editing. E.S.N.: Data collection, transcription, data curation, writing – review and editing. All authors read and approved the final manuscript.

## Data Availability

This study is based on in-depth qualitative interviews conducted in a single district-level hospital. Full interview transcripts cannot be shared publicly due to confidentiality risks and ethical restrictions, as participants could be identifiable from contextual details. Relevant data excerpts are included in the manuscript, and additional information may be made available upon reasonable request, subject to ethical approval.

## Acknowledgements

The authors would like to thank the management and staff of the study hospital for their cooperation and support. We are also grateful to all healthcare professionals who generously shared their time and experiences.

